# Using time use diaries to track changing behavior across successive stages of COVID-19 social restrictions

**DOI:** 10.1101/2021.01.29.21250766

**Authors:** Oriel Sullivan, Jonathan Gershuny, Almudena Sevilla, Francesca Foliano, Margarita Vega-Rapun, Juana Lamote de Grignon, Teresa Harms, Pierre Walthery

**Affiliations:** ESRC Centre for Time Use Research, Social Research Institute, University College London

**Keywords:** COVID-19 behavioral risk, time use diary methodology, comparison of lockdown behavior

## Abstract

How did people change their behavior over the different phases of the UK COVID-19 restrictions, and how did these changes affect their risk of being exposed to infection? Time use diary surveys are unique in providing a complete chronicle of daily behavior; 24-hour continuous records of the populations’ activities, their social context and their location. We present results from four such surveys, collected in real time from representative UK samples, both before, and at three points over the course of the current pandemic. Comparing across the four waves, we find evidence of substantial changes in the UK population’s behavior relating to activities, locations and social context. We assign different levels of risk to combinations of activities, locations and copresence, to compare risk-related behavior across successive ‘lockdowns’. We find evidence that during the second lockdown (November 2020) there was an increase in high-risk behaviors relative to the first (starting March 2020). This increase is shown to be associated with more paid work time in the workplace. At a time when capacity is still limited both in respect of immunization and track-trace technology, governments must continue to rely on changes in people’s daily behaviors to contain the spread of COVID-19 and similar viruses. Time use diary information of this type, collected in real time across the course of the COVID-19 pandemic, can provide policy-makers with information to assess and quantify changes in daily behaviors, and the impact they are likely to have on overall behavioral-associated risks.

## Introduction

How did people change their behavior over the different phases of the UK COVID-19 regulations on activities and social contact, and how did these changes affect their risk of being exposed to infection? Time use diary surveys are unique in providing a complete chronicle of daily behavior; 24-hour continuous records of the populations’ activities, their social context and their location. We present results from 4 such surveys, collected in real time from representative UK samples, both before, and at three points over the course of the current pandemic. Comparing across the four waves, we find evidence of significant changes in the UK population’s activities, locations and social interactions. Drawing on the epidemiological literature, we assign different levels of risk to specific combinations of activities, locations and copresence to show how these changes affected the populations’ risk of exposure to infection over the course of the pandemic. One of our main motivations is to compare behavior across successive ‘lockdowns’. We find evidence that the second lockdown (November 2020) was characterized by rather more high-risk behavior than the first (starting March 2020). We discuss possible reasons for this, drawing on research that distinguishes responses to differences in regulations from responses motivated by concern about changing rates of infectious transmission. At a time when capacity is still limited in respect both of immunization and track-trace technology, governments must continue to rely on changes in people’s daily behaviors to contain the spread of COVID-19 and similar viruses. Time use diary information of this type, collected in real time across the course of the pandemic, can provide policy-makers with information to assess and quantify the changes in daily behaviors that are associated with different phases of the COVID-19 pandemic, and the impact they are likely to have on overall behavioral-associated risks.

Complete spatio-temporal accounts of the activities (including their location and social context) and socio-demographic characteristics of representative samples are key to understanding populations’ changing behavioral risks of infection [1]. Epidemiological surveys of behavior during the pandemic have focused on measuring social contacts in order to determine risk [2,3,4,5], while social science surveys have mainly focused on asking respondents to estimate the quantity of time they spent in different activities at particular times [6]. Yet neither of these sources provides a complete record of daily behavior. Time use diaries have been used before in the context of investigating behavior related to COVID-19 [7,8,9], but not to report changing behavioral-related infection risks at successive periods of changing social restrictions (the UK Office of National Statistics collected pilot online time-use diaries in March-April 2020 and September-October 2020 [10], but did not take full advantage of the diary design by combining multiple diary fields of the diary to estimate behavior-related infection risks).

It is clear from the epidemiological literature that infection risk involves proximity to infected individuals. From our data, we assign levels of risk associated with daily behavior to combinations of activity, location and social context. When time use diary-derived patterns of daily behavior are linked to infection risk in this way, it enables the identification of those changes in behavior which are most likely to contribute to subsequent changes in infection rates. In respect of our main research question, the short second (November 2020) lockdown was associated with higher levels of high-risk daily behavior than the first (starting in March 2020). We disaggregate these changes to examine potential differentials by separate activity/copresence/location combinations, and by two characteristics known to be associated with the risk of infection; gender and age [11,12].

## Institutional context

Before proceeding, it is important to know something about the institutional context associated with the different phases of the COVID-19 crisis in the UK. In a previous article [1], we showed substantial differences between the 2016, pre-pandemic, distribution of risk-related activities and that for the first UK lockdown starting in late March 2020. In general, population-level patterns in time use change only gradually [13], so these previously-documented differences in time use patterns can with some confidence be attributed to a significant shift in behavior in response to the initial external shock of the pandemic. The main focus of this paper is a comparison of risk-related behavior during the first lockdown (starting March 2020) with such behavior in the second lockdown (starting November 2020). Comparing these two periods, there was little difference in regulations in England (the country of 87% of all survey respondents^1^) between the first and second lockdowns in terms of restrictions on social gathering, exercise, the hospitality sector and business opening [14]. The underlying rule remained ‘stay at home’. Essential workers were permitted to leave home to work, but non-essential businesses including hospitality had to close or operate deliveries/takeaways only. The main difference in regulations was that during the second lockdown schools remained open, while regulations on social gathering were more restrictive than during the later part of the first lockdown (from June, people were allowed to meet outdoors with up to 6 people, instead of with just one for exercise in November). However, in the period of relaxation of restrictions between lockdowns, in August 2020, gatherings of up to thirty people were legally permitted, indoors and out, and many businesses re-opened (although with social distancing restrictions in place). The second lockdown was imposed on 5 November, following a period of tightening of rules that began in mid-September, in response to rapidly rising infection and death rates (the ‘second wave’ of UK COVID-19 infections).

## Data

The data were collected via a low-respondent-burden (12-15 minutes on average per day), online time use diary instrument (the Click and Drag Diary Instrument, CaDDI), suitable for rapid deployment in real time in situations such as the current pandemic, and described in detail in a previous article [15]. Information was collected on six characteristics (or ‘diary fields’): ‘main’ and ‘other simultaneous’ activities; location; co-presence; ICT device use; and enjoyment, for each successive episode throughout the 24-hour day (where ‘episodes’ are defined as periods through which all diary fields remain unchanged). Continuous diary accounts recording successive activities are regarded as superior in the measurement of daily behavior to survey questions about the frequency or duration of activities, because they reduce recall issues (being generally recorded on the diary day as a continuous sequence of activities, aiding recall), deter misrepresentation (since the sequence format does not lend itself so easily to under- or over-representation of particular activities), and enhance reliability (as different durations of the same activity occurring throughout the day may be recorded). Time use diary data have been validated through small-scale comparisons with more expensive approaches using objective instruments (cameras, motion sensors) worn through the diary observation period [16].

Respondents were members of the large Dynata agency market research panel, who volunteered for the surveys and were selected on the basis of age, sex, social grade and region quotas that were nationally representative of the 2016 population. The four cross-sectional sample waves were collected, respectively, in: February, October and December (to reduce single-season effects) of 2016 (representing pre-pandemic behavior patterns, N=1011 diary days); May-June 2020 (during the first UK lockdown, N=1007); August 2020 (during the post-lockdown summer relaxation of restrictions, N=987); and November 2020 (the second lockdown, N=1358). Each respondent completed diaries for between 1 and 3 days, yielding a total across the four waves of the survey of 4360 days from 2202 individuals. Weights were calculated to yield the correct mix of days-of-the-week for each sex by (10-year) age group, and to correct for the 2016 age group quotas within waves. All analyses in this article were conducted using these weights. A more detailed description of the quota distributions across waves is found in Gershuny et. al. 2021 [1]. The data are available from the core collection of the UK Data Archive, Study number 8741.

Because of the paucity of real-time representative data, particularly at the start of the pandemic, social scientists have often relied on commercially-run panels to understand the outcomes of the COVID crises, due to their rapid response times [17, 18]. The CaDDI sample merits some claim to representativeness as it is based on nationally representative quotas for age-group, sex, region and social group in 2016. As a robustness check, Supporting Information Table S1 shows a comparison of mean minutes spent in the CaDDI high and low-risk categories (used in our main regression analysis of Table 2) with the nationally representative 2014-15 UK Time Use Survey (UKTUS) of 2014-15. Time spent in high-risk behaviors (i.e. high-risk activity/location and copresence combinations), the main focus of our analysis, aligns well between the two surveys: the difference is small, at 3 minutes/day on average, and is not statistically significant. Respondents to CaDDI recorded somewhat longer time spent in low-risk behaviors in 2016 than respondents in the UKTUS (a difference of 26 minutes/day, P<.05). It may be that there is some respondent bias, with those spending longer at home also being more likely to complete CaDDI-type surveys, but there could also have been other differences relating to the timing of the CaDDI survey (which took place in February, October and December of 2016 as opposed to across a full year in 2014-15).

**Table 1.**
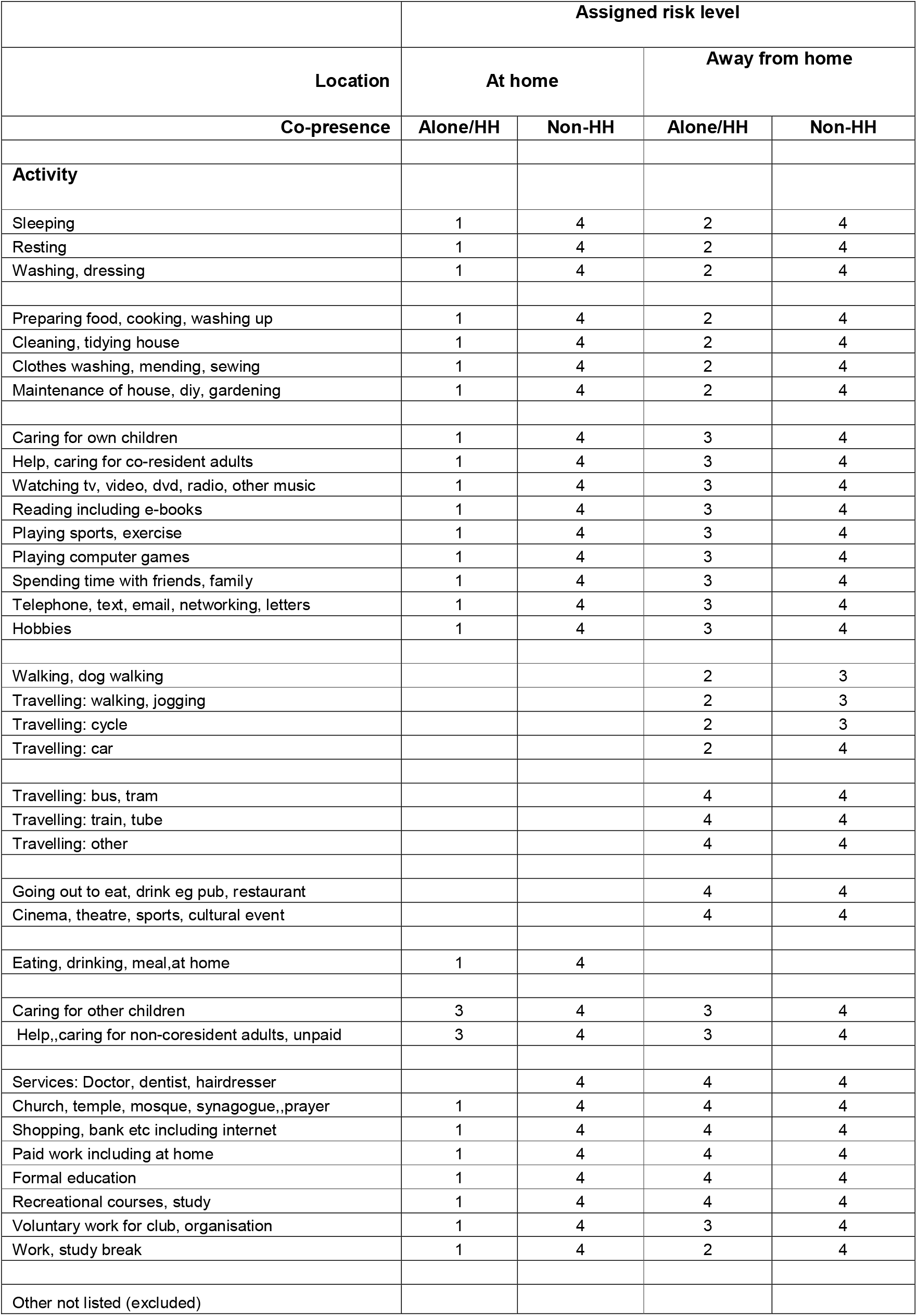
Risk-level assignments, by activity, location and co-presence combined categories.

|  | Assigned risk level |  |  |  |
| --- | --- | --- | --- | --- |
| Location | At home |  | Away from home |  |
| Co-presence | Alone/HH | Non-HH | Alone/HH | Non-HH |
| Activity |  |  |  |  |
| Sleeping | 1 | 4 | 2 | 4 |
| Resting | 1 | 4 | 2 | 4 |
| Washing, dressing | 1 | 4 | 2 | 4 |
| Preparing food, cooking, washing up | 1 | 4 | 2 | 4 |
| Cleaning, tidying house | 1 | 4 | 2 | 4 |
| Clothes washing, mending, sewing | 1 | 4 | 2 | 4 |
| Maintenance of house, diy, gardening | 1 | 4 | 2 | 4 |
| Caring for own children | 1 | 4 | 3 | 4 |
| Help, caring for co-resident adults | 1 | 4 | 3 | 4 |
| Watching tv, video, dvd, radio, other music | 1 | 4 | 3 | 4 |
| Reading including e-books | 1 | 4 | 3 | 4 |
| Playing sports, exercise | 1 | 4 | 3 | 4 |
| Playing computer games | 1 | 4 | 3 | 4 |
| Spending time with friends, family | 1 | 4 | 3 | 4 |
| Telephone, text, email, networking, letters | 1 | 4 | 3 | 4 |
| Hobbies | 1 | 4 | 3 | 4 |
| Walking, dog walking |  |  | 2 | 3 |
| Travelling: walking, jogging |  |  | 2 | 3 |
| Travelling: cycle |  |  | 2 | 3 |
| Travelling: car |  |  | 2 | 4 |
| Travelling: bus, tram |  |  | 4 | 4 |
| Travelling: train, tube |  |  | 4 | 4 |
| Travelling: other |  |  | 4 | 4 |
| Going out to eat, drink eg pub, restaurant |  |  | 4 | 4 |
| Cinema, theatre, sports, cultural event |  |  | 4 | 4 |
| Eating, drinking, meal,at home | 1 | 4 |  |  |
| Caring for other children | 3 | 4 | 3 | 4 |
| Help,,caring for non-coresident adults, unpaid | 3 | 4 | 3 | 4 |
| Services: Doctor, dentist, hairdresser |  | 4 | 4 | 4 |
| Church, temple, mosque, synagogue,,prayer | 1 | 4 | 4 | 4 |
| Shopping, bank etc including internet | 1 | 4 | 4 | 4 |
| Paid work including at home | 1 | 4 | 4 | 4 |
| Formal education | 1 | 4 | 4 | 4 |
| Recreational courses, study | 1 | 4 | 4 | 4 |
| Voluntary work for club, organisation | 1 | 4 | 3 | 4 |
| Work, study break | 1 | 4 | 2 | 4 |
| Other not listed (excluded) |  |  |  |  |

**Table 2.** Changes in high and low activity risk levels across successive stages of COVID-19 social restrictions: *OLS Regression coefficients, minutes per day: N=4360 days, clustered by 2202 respondents*

|  | **Risk level 4**<br>**(high-risk)** | **Risk level 1**<br>**(low-risk)** |
| --- | --- | --- |
| **Survey dates (Regulatory phases)** |  |  |
| March & Oct 2016 (Pre COVID-19) | 195*** | -233*** |
| May-June '20 (1<sup>st</sup> Lockdown) | (ref.) | (ref.) |
| August '20 (Relaxation) | 65*** | -88*** |
| November '20 (2<sup>nd</sup> Lockdown) | 35** | -29 |
| **Sex** |  |  |
| woman | -7 | 16 |
| man | (ref.) | (ref.) |
| **Age group** |  |  |
| aged 18-24 | 56** | -86** |
| aged 25-34 | 82*** | -115*** |
| aged 35-44 | 102*** | -128*** |
| aged 45-54 | 83*** | -91*** |
| aged 55-64 | 56*** | -89*** |
| aged 65+ | (ref.) | (ref.) |
| **Social grade** |  |  |
| A,B | -12 | -13 |
| C1, C2 | (ref.) | (ref.) |
| D,E | -55*** | 55*** |
| **Constant** | 53*** | 1333*** |
| **R<sup>2</sup>** | 0.12 | 0.11 |

Key: \*\*\* P<.001, \*\* P<.01, \* P<.05

## Methods

We assigned the level of risk for each diary episode based on combinations of three of the simultaneous diary fields: activity type, location and co-presence. These assignments are made taking cognizance of the literature on COVID-19 infection transmission, which considers time at home alone or with members of the same household as lowest-risk, with the main focus for transmission being contact with non-household members, both at, or away from, home. The virus is more likely to be transmitted indoors, in unventilated spaces, in crowds, and through prolonged personal contact [19]. Table 1 shows the detailed assignments for each combination of the three diary fields to one of four risk categories, ranging from lowest (1) to highest risk level (4)^2^. Activities are shown down the first column, and are assigned a risk category according to their combinations with recorded co-presence (‘alone or with other household members’/’with non-household members’), and location (‘at home’/’away from home’). Estimates of risk vary according to the activity (e.g. cinema implies the presence of other, non-household, individuals), and are also influenced by its characteristic location (e.g. indoors enclosed, vs open-air). While co-presence information has been shown to be subject to non-response and discrepancies of record between spouses in some time use diary surveys [20] the CaDDI instrument mitigates the non-response issue by requiring the completion of co-presence information before respondents can continue the diary. To the extent that any remaining measurement error in the risk categories associated with the co-presence variable does not change over the period of the CaDDI waves in a systematic way, our estimated coefficients for the risk category variables will remain unbiased. In addition, information on co-presence is supplemented from the activity fields in the attribution of risk, so, for example ‘using public transport’ is taken to imply current presence of other, non-household, individuals. Finally, to take exposure duration into account we assigned all activity combinations lasting only one 10-minute timeslot to the lowest risk level (level 1) [21]. For a more detailed description of these risk assignments and their rationale see Table 1 and Gershuny et al 2020 [1].^3^

We used multivariate OLS regression models to investigate the statistical significance of differences in the time spent in high and low risk categories across the waves of the data. In these models the dependent variable is the time spent at each wave in the high (level 4) and low (level 1) risk categories, and the independent variables are survey wave, gender, age group and social grade. To estimate statistical significance we used robust clustered standard errors estimated from single-respondent cross-day clusters (Stata vs 16), to take account of the varying number of diaries per respondent.

## Findings

Figure 1 illustrates descriptive changes in behavior across different combinations of activity, location and co-presence for the 4 survey waves. It summarizes the ‘average day’ (1440 minutes) for each wave as mean daily durations (minutes per day) disaggregated into 9 different combinations of activity, location, and co-presence (plus one small unallocated category). Onto this breakdown of activity combinations we have superimposed the 4 risk levels shown in Table 1. Changes across the four columns of Figure 1 are larger than those observed in the UK population’s time allocation over the fifteen-year period 2000-2015^4^. The initial lockdown period (data collected in May-June 2020) was associated with a substantial shift of time away from the high-risk behaviors characterizing pre-pandemic behavior (in particular, paid work in the workplace and out-of-home leisure) towards home-based activities involving lower levels of contact with non-household members (with lower risk of infection). Partial relaxation of the lockdown regulations in August 2020 was associated with the expected partial return to previous patterns of daily activity. Finally, the subsequent re-imposition of lockdown regulations during November 2020, as we show below, was associated with a return to the patterns of behavior observed during the first lockdown, with some important differences.

**Figure 1:**
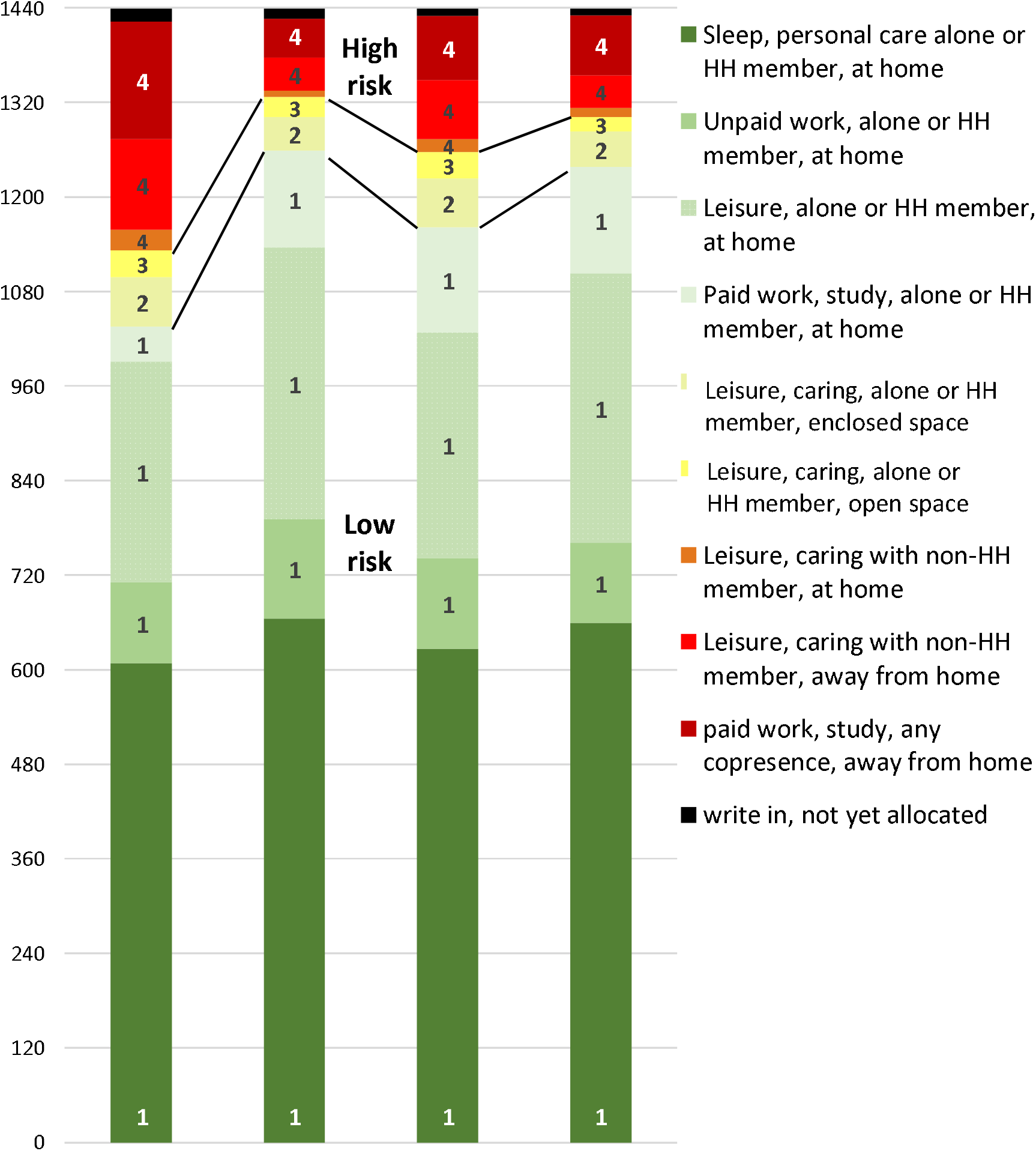
Changes in UK time use (minutes per day) by level of risk of activity combinations a cross successive phases of COVID-19 social restrictions. Notes: May ‘20 = first lockdown Nov ‘20 = second lockdown Risk category 1= low-risk; risk category 4=high-risk ‘Write-in, not yet allocated’ represents the small proportion of time as yet uncoded

Were these changes statistically significant? Table 2 shows results from an OLS regression model, using clustered standard errors. Two models are shown, with time (mean minutes/day) spent in high and low-risk levels (levels 4 and 1) respectively as the dependent variable. The models include dummy variables for survey wave, sex, age group and social grade. The models for high and low-risk activities account for 12 and 11% of the explained variance (R^2^) respectively, and they provide support for the straightforward reading of Figure 1. The substantial reduction of time per day in high-risk, and increase in time in low-risk, activity combinations associated with the movement into the initial lockdown are both highly statistically significant. Holding other variables constant, prior to the pandemic in 2016, an average of 195 more minutes per day were spent in high-risk activities and 233 fewer minutes in low-risk activities compared to first lockdown (both differences at P<.001). Differences in the opposite direction, between May-June 2020 (1^st^ lockdown) and August 2020 (relaxation of restrictions) are again strongly statistically significant, though without returning to previous (pre-lockdown) levels of riskiness. During this intermediate period there was an average of 65 more minutes per day spent in high-risk activities and 88 minutes less in low-risk activities (both P<.001) compared to first lockdown; about a third of the size of the change between pre-pandemic levels and first lockdown. Of particular interest for our research question is that the return to lockdown in November 2020 was associated with higher levels of high-risk behavior than during first lockdown, with 35 more minutes per day spent in high-risk activities (P<.01). In order to understand what activity combinations were associated with these differences, the following section describes differences in activity/copresence/location combinations across the survey waves.

### Disaggregation of cross-wave changes by activity/co-presence/location categories

Table 3 provides more detail on the changes in behavior that are associated with these differences in risk levels, again expressed as contrasts with behavior during the first lockdown. Nine OLS models are shown across the rows of the table, each showing regression coefficients for the mean minutes/day spent in each of the nine activity/co-presence/location categories shown in Figure 1. Each model includes the same list of control variables as the models shown for Table 2 (see Supporting Information Table S2 for the full set of coefficients from the regression models).

**Table 3.** Changes in 9 activity combinations across successive stages of COVID-19 social restrictions: *OLS Regression coefficients, minutes per day: N=4360 days, clustered by 2202 respondents*

| Activity combination category | Activity/copresence/location combination | Risk level | 2016 | May-Jun'20 | Aug '20 | Nov '20 |
| --- | --- | --- | --- | --- | --- | --- |
| 1 | Sleep, personal care, alone or with HH member, at home | 1 | -62 *** | (ref.) | -34** | -9 |
| 2 | Unpaid work, alone or with HH member, at home | 1 | -28*** | (ref.) | -14 | -30*** |
| 3 | Leisure, alone or with HH member, at home | 1 | -75*** | (ref.) | -45** | -11 |
| 4 | Paid work, study, alone/HH member, at home | 1 | -68*** | (ref.) | 5 | 21* |
| 5 | Leisure, caring, alone/with HH member, away, open space | 2 | 23*** | (ref.) | 18** | 4 |
| 6 | Leisure/caring, alone/with HH member, away, enclosed space | 3 | 11* | (ref.) | 8 | -6 |
| 7 | Leisure/caring or paid work with non-HH member at home | 4 | 18*** | (ref.) | 7 | 4 |
| 8 | Leisure/caring, with non-HH member, away from home | 4 | 71*** | (ref.) | 33*** | -2 |
| 9 | Paid work or study, any co-presence, away from home | 4 | 106*** | (ref.) | 25* | 33*** |

Key: \*\*\* P<.001, \*\* P<.01, \* P<.05

The predominantly negative coefficients of the first four rows of Table 3 indicate the higher levels of low-risk at-home activity combinations (including personal care, unpaid work, and home leisure) done during first lockdown. An exception was the greater time spent in paid work or study at home during the November lockdown. Correspondingly, less time was spent during first lockdown in activities away from home, and with people from outside the household (both at, and away from, home), in leisure or caring, and paid work or study (note statistically significant positive coefficients in rows 5 to 9).

All activity combinations were highly statistically significantly different in 2016 from the time spent in these activities at first lockdown, which saw reductions of 71 minutes per day in out-of-home leisure with non-household members (category 8), and a one-hour-45-minutes reduction in paid work/study away from home (category 9).The intermediate period (relaxation of restrictions in August 2020) produced, as Table 2 has shown, a partial return to these pre-COVID behavior patterns. In particular, less time was spent in personal care (including sleeping) at home, and at-home leisure alone or with household members (both at P<.01) than during first lockdown. There was no change, however, in time spent doing paid or unpaid work at home compared to first lockdown. At the other end of the scale, there was a half-hour average increase in the time spent in leisure or caring activities with non-household members outside of the home (category 8; P<.001). In addition, the high-risk return to the workplace (category 9) during this period of relaxation of restrictions, though statistically significant (P<.05), was only a quarter of the size of the previous shift away from paid work at the workplace during first lockdown (25 minutes more on average, compared to one-hour-and-45-minutes less).

Table 2 showed that the second, November, lockdown involved a return to a less risky behavior pattern, similar to that characterizing the shift into first lockdown, but with some interesting differences. The second lockdown was characterized by half an hour less time spent doing unpaid work at home (category 2; P<.001), but more time spent doing paid work at home, which has higher by 21 minutes on average than during first lockdown (P<.05). Table 2 also showed that there was a greater amount of time spent in high-risk, out of home, activities during the second lockdown compared to the first. In Table 3 this is shown to be associated with time spent doing paid work in the workplace - over half an hour more than during first lockdown (33 minutes, P<.001), a difference greater than, but more similar to, that during the August relaxation of restrictions (25 minutes, P<.05) than to first lockdown.

### Disaggregation of cross-wave changes by age and gender

Age and gender are both factors which have been shown to be associated with important differences in infection risks [11,12]; do the differences shown in Table 2 apply across these subgroups? Figure 2 plots regression coefficients for time spent (predicted mean minutes per day) in the high and low-risk activity combinations (levels 4 and 1 respectively), expressed as contrasts to first lockdown, for both genders (respondents were asked to report their biological sex) and three age groups. For these analyses ages were combined into three groups due to sample number restrictions, which also meant that it was not possible to analyze age groups by gender. The same combination of dependent and independent variables is used as in the regression models for Table 2 (omitting the gender variable in the analyses disaggregated by gender, and the age group variable in the analyses disaggregated by age group). Due to sample number constraints we also omit the gender variable and combine social grade into 2 categories for the age group analyses. Full regression coefficients are provided in Supporting Information Tables S3 (by gender) and S4 (by age group).

**Figure 2.**
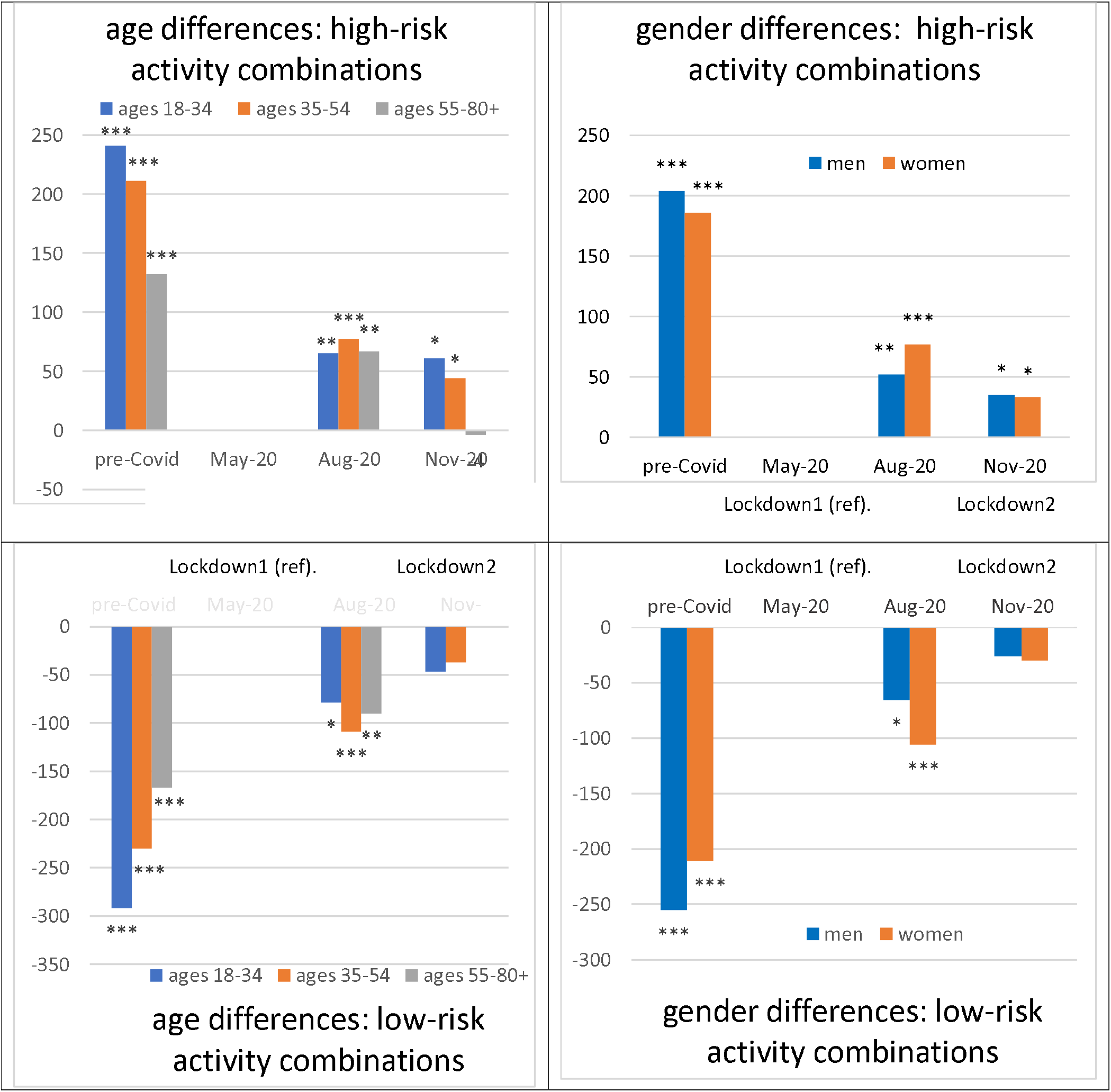
Age and gender differences across successive stages of COVID-19 social restrictions: *OLS Regression coefficients, minutes/day* Key: *** P<.001, ** P<.01, * P<.05 Note: Full model results are available in the Supporting Information, Tables S3 and S4

The pattern of the coefficients across the stages of social restrictions shown in Figure 2 broadly reflects the overall pattern shown in Table 2. For high-risk activity combinations (upper pair of charts), there were substantial and strongly significant differences pre-pandemic (more high-risk time) compared to the first lockdown (less high-risk time) across all age groups and both genders. During August 2020 there was a less dramatic reversal, with more time again being spent in high-risk activity combinations, again for both genders and all age groups. Women spent on average rather more time than men in high-risk activity combinations during the relaxation of restrictions in August 2020 (by 25 minutes/day). The amount of time spent in high-risk activity combinations during the second (November) lockdown was significantly greater than during first lockdown for both genders (by 35 and 33 minutes/day; both P<.05), and for the two younger age groups (61 minutes/day in the case of 18-34 year olds, and 44 minutes in the case of 35-54 year olds; both P<.05). However, there was no statistically significant difference between lockdowns for the oldest age group (aged 55+).

The lower pair of graphs show the time spent in low-risk activity combinations compared to first lockdown; coefficients here are shown on a negative scale. In accordance with the results of Table 2 there was an initial very substantial increase in low-risk at-home activities between the pre-pandemic period and first lockdown across both genders and all age groups. Again, lockdown was followed by a smaller reversal away from these low-risk activity combinations, particularly for women, and among those aged 35-54. There was no statistically significant difference in the time spent in these low risk activity combinations between first and second lockdown.

## Discussion

We show changes in behavior across the phases of the COVID-19 pandemic in the UK, collected in real time at 3 time-points characterized by different regulations, and a 2016 pre-pandemic baseline (illustrated in Figure 1). Assigning risk levels to different combinations of activities, location and copresence from the diary fields, we show significant differences between the populations’ behavior during lockdowns (in May-June 2020 and November 2020 – see Table 2). Both lockdowns were characterized by substantial increases in low-risk behavior compared both to the pre-pandemic baseline in 2016, and to the period of relaxation of restrictions in August 2020. These discontinuities in patterns of behavior within relatively short periods of time are unusual for trends in time use, which are generally characterized by gradual change, suggesting that they were a direct response to the external shock of the COVID-19 pandemic.

One of our primary motivations was to investigate if there were any differences in population behavior between the first and second lockdowns. We do indeed find evidence for more high-risk behavior during the November lockdown (see Table 2). Holding constant gender, age and social grade the population spent on average 35 more minutes per day in high-risk activities in late November 2020 than in first lockdown (May-June 2020). Looking in more detail at the types of activity/location/co-presence combinations engaged in (Table 3), the November lockdown was characterized by more time spent in paid work at the workplace (33 minutes/day, P<.001). Disaggregating the changes by gender and age group shows that these differences in time spent in high-risk activities between the two lockdown periods applied across all subgroups, with the exception of the oldest age group, aged 55+, whose high-risk behavior during the second lockdown was not statistically significantly different from that during first lockdown (Figure 2, upper pair of graphs).

How can we account for the associations we have found between changing risk-related behaviors and the varying phases of the COVID-19 pandemic? In particular, the fact that there was more time spent in high-risk activity combinations in the second, compared to the first, lockdown? Other research has identified possible mechanisms (not necessarily mutually exclusive) that might be involved in explaining such differences: 1) regulations associated with lockdown were different; 2) perceptions of risk were different; 3) people were tired/resentful of continuing restrictions. Mendola et al., in a recent study comparing human mobility responses to changes in government regulations with responses to wider perceptions of risk based on information about infection transmission, concluded that responses to government-imposed regulations had a stronger effect than information about risk [24]. We cannot directly adjudicate between these hypotheses using our data; however, to be persuasive, such hypotheses must be capable of accounting for the key aspects of the evidence that we highlight here. The first is the timing: we have shown that time spent in high-risk activity/location/copresence combinations increased in the inter-lockdown period, and was somewhat higher during the second lockdown than the first. The second is the composition: the greater amount of time spent in high-risk behavior during the second lockdown was associated with more time spent in paid work in the workplace. We find no evidence for behavioral ‘fatigue’ with regulations, as we observe no more time being spent in out-of-home leisure-and-caring-related activity combinations during the second lockdown than during the first. In relation to perceptions of risk, the second UK lockdown (November 2020) was imposed in direct response to rapidly rising infection and death rates (the ‘second COVID-19 wave’) occurring from mid-September 2020 onwards as a result of the loosening of restrictions during August [25]. The national context suggests that people were well aware of the reasons for the urgent imposition of the second lockdown, which were extensively reported in the media [26]. Nevertheless, we find an increase in high-risk behavior during second lockdown (with the exception of the age group aged 55 and over, less likely to be in paid employment, and who may also have been more cautious due to awareness of age-related risk). In order to account for this increase we suggest that the fact that schools were open during the second lockdown may have enabled some parents to return to the workplace (i.e. a regulatory difference). Also consistent with the increased amount of time spent at the workplace that we document for the second lockdown is that, despite the fact that regulations governing business and hospitality were the same, more businesses were open during the second lockdown than during the first (operating within lockdown rules - for example, by providing takeaway food) following a period of relaxation of restrictions in which more general opening was permitted [27].

At a time when capacity is still limited both in respect both of immunization and track-trace technology, governments must continue to rely on changes in people’s daily behaviors to contain the spread of COVID-19 and similar viruses. The evidence presented here should help to increase policy awareness of the extent to which risk-related population behavior may be expected to change in response to changing infection risks and associated regulations during the continuation of the COVID-19 crisis, and in any future pandemic with similar characteristics. Lockdowns, and the reasons for them, do have a significant effect on population behavior, but some lockdowns appear to be more strongly associated with high-risk behavior than others. In particular, the fact that schools were open in a context where more businesses also remained open likely contributed to the greater amount of high-risk time spent in the workplace during the second lockdown. This detailed analysis of successive UK lockdowns throws light on what the consequences of different policy approaches might be to changing risk-related behavior. In future research such data, sampled randomly from a national population frame, and more frequently–either monthly, or, preferably, continuously—could significantly enhance policy understanding of such changes through enabling the tracking of risk-related behavior directly alongside changes in rates of infection.

## Data Availability

The data is available from the UK Data Service archive

https://beta.ukdataservice.ac.uk/datacatalogue/studies/study?id=8741

## Acknowledgements

the writing of this paper was funded by the Economic and Social Research Council (ES/S010149/1), and the European Research Council (FP7 770839).

## Supplementary Information

**Table S1.** Comparison of CaDDI and UK Time Use Survey (UKTUS) of 2014-15 (high and low-risk levels)

|  | **CaDDI 2016**<br>**(mins/day)** | **UKTUS 2014-15**<br>**(mins/day)** | **Difference** | **t** | **P-value** |
| --- | --- | --- | --- | --- | --- |
| **Low-risk (level 1)** | 1026 *(314)* | 1000 *(333)* | 26 | 2.13 | 0.03 |
| **High-risk (level 4)** | 302 *(267)* | 299 *(262)* | 3 | 0.32 | 0.75 |
| **N** | 1011 | 7656 |  |  |  |

Notes: This table compares the mean daily minutes spent in low and high-risk levels in CaDDI 2016 and UKTUS 2014-15.
Bracketed italics show standard deviations around the means
The sample of UKTUS diaries included in the comparison is restricted to those diaries with less than 30 mins missing information on activities, copresence or location.

**Table S2.** Full models for 9 activity combinations. (1=lowest risk, 9=highest): OLS Regression coefficients, minutes per day *(N=4360 days, 2202 respondents)*

| Activity combinations | 1. Sleep etc, alone/HH, at home | 2.Unpaid etc, alone/HH, at home | 3.Leisure etc, alone/HH, at home | 4. Paid etc., alone/HH, at home | 5.Leisure etc, alone/HH, open space | 6.Leisure etc, alone/HH, enclosed | 7.Leisure etc, non-HH, at home | 8.Leisure etc, non-HH, away | 9. Paid etc, any co-pres, away |
| --- | --- | --- | --- | --- | --- | --- | --- | --- | --- |
| 2016 | -62*** | -28*** | -75*** | -68*** | 23*** | 11** | 18*** | 71*** | 106*** |
| May-20 | (ref.) | (ref.) | (ref.) | (ref.) | (ref.) | (ref.) | (ref.) | (ref.) | (ref.) |
| Aug-20 | -34*** | -14* | -45*** | 5 | 18*** | 8 | 7* | 33*** | 25** |
| Nov-20 | -9 | -30*** | -11 | 21** | 4 | -6 | 4 | -2 | 33*** |
| woman | 4 | 51*** | -32*** | -7 | -6 | -1 | 8** | 14*** | -29*** |
| man | (ref.) | (ref.) | (ref.) | (ref.) | (ref.) | (ref.) | (ref.) | (ref.) | (ref.) |
| aged 18-24 | -22 | -90*** | -79*** | 104*** | 13 | 22** | 2 | -37*** | 90*** |
| aged 25-34 | -36** | -25** | -159*** | 105*** | 26*** | 13** | 11 | -25*** | 96*** |
| aged 35-44 | -56*** | -9 | -149*** | 85*** | 17*** | 13*** | 5 | -14* | 111*** |
| aged 45-54 | -49*** | -25** | -88*** | 71*** | 7 | 3 | 6 | -18** | 95*** |
| aged 55-64 | -37*** | -12 | -78*** | 38*** | 22*** | 7 | 4 | -8 | 60*** |
| aged 65+ | (ref.) | (ref.) | (ref.) | (ref.) | (ref.) | (ref.) | (ref.) | (ref.) | (ref.) |
| Soc gr. AB | -17* | -2 | -31*** | 37*** | 17*** | 11*** | -2 | 3 | -14 |
| C1,C2 | (ref.) | (ref.) | (ref.) | (ref.) | (ref.) | (ref.) | (ref.) | (ref.) | (ref.) |
| DE | 17 | 42*** | 52*** | -56*** | -7* | 2 | 8 | -10* | -53*** |
| (Constant) | 702*** | 116*** | 458*** | 57*** | 26*** | 12*** | -2 | 53*** | 2 |
| RSq | 0.025 | 0.081 | 0.096 | 0.100 | 0.019 | 0.014 | 0.010 | 0.069 | 0.092 |

Key: \*\*\* P<.001, \*\* P<.01, \* P<.05

**Table S3.** Full models for high and low risk activity categories modelled by gender: OLS Regression coefficients: minutes per day *(N=4360 days, 2202 respondents)*

| **Women** | **High risk activities**<br>**(level 4)** | **Low risk activities**<br>**(level 1)** |
| --- | --- | --- |
| 2016 | 186*** | -212*** |
| Aug 2020 | 77*** | -106*** |
| Nov 2020 | 33** | -30 |
| Aged 18-24 | 46** | -71** |
| Aged 25-34 | 58*** | -60** |
| Aged 35-44 | 53*** | -69*** |
| Aged 45-54 | 75*** | -80*** |
| Aged 55-64 | 48** | -79*** |
| Social grade AB | 3 | -21 |
| Social grade DE | -48*** | 48*** |
| Model constant | 57*** | 1,325*** |
| R-squared | 0.108 | 0.092 |
| **Men** |  |  |
| 2016 | 204*** | -255*** |
| Aug 2020 | 52*** | -66** |
| Nov 2020 | 35** | -26 |
| Aged 18-24 | 54 | -77 |
| Aged 25-34 | 103*** | -166*** |
| Aged 35-44 | 146*** | -182*** |
| Aged 45-54 | 90*** | -99*** |
| Aged 55-64 | 63*** | -95*** |
| Social grade AB | -21 | -13 |
| Social grade DE | -57*** | 56** |
| Model constant | 41** | 1,355*** |
| R-squared | 0.133 | 0.128 |

**Table S4.** Full models for high and low risk activity categories modelled by age-group: OLS Regression coefficients, minutes per day (*N=4360 days, 2202 respondents)*

| **High-risk activities (level 4)** | **ages 18-34** | **ages 35-54** | **ages 55-80+** |
| --- | --- | --- | --- |
| CaDDI 2016 | 241*** | 211*** | 132*** |
| CaDDI Aug 2020 | 65*** | 77*** | 67*** |
| CaDDI Nov 2020 | 61** | 44** | -4 |
| Social grade DE | -51** | -57*** | -43*** |
| Constant | 100*** | 129*** | 101*** |
| R-Squared | 0.129 | 0.093 | 0.094 |
| **Low-risk activities (level 1)** |  |  |  |
| CaDDI 2016 | -292*** | -230*** | -167*** |
| CaDDI Aug 2020 | -79* | -109*** | -90*** |
| CaDDI Nov 2020 | -47 | -37* | 8 |
| Social grade DE | 70* | 69*** | 53*** |
| Constant | 1,247*** | 1,230*** | 1,258*** |
| R-Squared | 0.113 | 0.081 | 0.087 |

## Footnotes

1 The quota-based selection of respondents by country/region reflects the geographical population distribution of the UK. Although there were some differences in specific restrictions and their timings between countries (as well as between areas of England at certain times), the basic parameters across the pandemic period of: lockdown1; relaxations of restrictions; and restriction tightening (lockdown 2 in England), were the same over the period covered by the three CaDDI ‘pandemic’ surveys.

2 Ultimately, the number of risk categories used is an empirical question that cannot be resolved purely on theoretical grounds, and depends on the evidence and data available. For instance the Infectious Diseases Society of America (IDSA) uses a 3-way classification based on less detailed behavioral information than are available in the CaDDI time use diary surveys: *https://www.idsociety.org/globalassets/idsa/public-health/covid-19/activity-risk.pdf*

3 Recent developments in understanding of the transmission of the virus means that we have for this article updated our scale to take account of increasing evidence from the epidemiological literature for the importance of ventilation in the risk of infection transmission [22, 23], now also assigning all those activities that take place at home but in the presence of non-household others to the highest risk category (level 4).

4 Refer to Chapter 1 of reference [13]

